# Outcomes of COVID-19 infection in patients with chronic kidney disease on maintenance hemodialysis at a COVID hospital in India

**DOI:** 10.1101/2022.01.07.22268915

**Authors:** Ram Singh, Sudarsan Krishnasamy, Jitendra Kr Meena, Prashant Sirohiya, Balbir Kumar, Brajesh kumar Ratre, Saurabh Vig, Anuja Pandit, Hari Sagiraju, Raghav Gupta, Sushma Bhatnagar

## Abstract

**Background:** Patients with chronic kidney disease (CKD) on hemodialysis are highly vulnerable to COVID-19 infection with a mortality rate higher than the rest of the population. There are several clinical and laboratory parameters that can predict the course and the outcomes in this group of population.

**Methods:** We retrospectively collected the baseline demographic, clinical, in-hospital, and laboratory data of the patients with CKD on maintenance hemodialysis who were admitted to our COVID-19 hospital during the first and the second wave.

**Results:** We obtained data for 35 patients from the first and 5 patients from the second wave. The analysis of the data for 35 patients from the first wave revealed shortness of breath (62.9%), and fever (54.3%) being the most common presenting symptoms, and the majority of the patients (57.2 %) presented with moderate to severe disease at admission with 57 % had bilateral lung infiltrates, and required oxygen support (65.7%) at admission. The comparison of clinical and laboratory markers between survivors (27 patients, 77.1%) and non-survivors (8 patients, 22.9%) revealed an older age, severe disease at presentation, invasive mechanical ventilation, baseline severe lymphocytopenia, high serum glutamic oxaloacetic transaminase, blood urea, and inflammatory markers like Interleukin-6 and procalcitonin, fibrinogen and low albumin in non survivors.

**Conclusions:** The older age, severe disease at presentation, the requirement of invasive mechanical ventilation, raised baseline Interleukin-6, procalcitonin, serum glutamic oxaloacetic transaminase, blood urea and a low albumin level could be valuable predictors of poor outcomes.

## Introduction

The course of Coronavirus disease 2019 (COVID-19) infection in patients with pre-existing kidney disease is quite severe and associated with high mortality as observed across the recent literature. A high vulnerability to infections had been observed in patients with chronic kidney disease (CKD) on maintenance hemodialysis(1) with an increased risk of severe disease and poor outcomes upon infection with COVID-19.(2) The dysregulation of immune response and coagulation abnormalities are postulated to be involved in the immunopathological process.(3) The older age, the burden of comorbidities, and frailty of CKD patients on maintenance dialysis together with inflammatory stress imposed by the infection pose high vulnerability to the worse outcomes.(4,5) The clinical characteristics and in-hospital course of CKD patients with COVID-19 infection and outcomes in terms of in-hospital mortality were studied retrospectively from the medical records. The comparison of clinical and laboratory parameters between survivors and non-survivors revealed a variety of factors that can affect and predict the clinical course of COVID-19 infection in these patients. We highlighted these factors in the Indian patients with CKD on maintenance hemodialysis who were admitted to the COVID-19 hospital at National Cancer Institute (NCI), All India Institute of Medical Sciences (AIIMS), Jhajjar during both the waves of COVID-19 from July 2020 to June 2021.

## Methods

This was a retrospective record review conducted at a tertiary care institute, National Cancer Institute (NCI), All India Institute of Medical Sciences (AIIMS) at Jhajjar, India. During the pandemic, the institute had been designated as a COVID hospital. The study protocol was approved by the Institute Ethics Committee of AIIMS, New Delhi. The protocol was designed and drafted following the Strengthening the Reporting of Observational studies in Epidemiology (STROBE) checklist. From the hospital electronic database, we included all consecutive patients who were known cases of CKD on maintenance hemodialysis with the existence of COVID-19 infection confirmed by the real-time polymerase chain reaction (RT-PCR) test from the nasopharyngeal swab sample. We excluded patients with inadequate data from the study.

We retrospectively collected the clinical data from the case files, screening forms, and treatment sheets. The laboratory parameters were abstracted into excel datasheet from the hospital electronic patient information portal. The demographic parameters like age, gender, comorbidities status, vital parameters including oxygen saturation at presentation, hemodialysis sessions, complication during or after the dialysis, medical treatment administered, course of disease during hospitalization, lab parameters at baseline, length of hospital stay and the outcomes in terms of discharge or death at end of hospitalization were collected.

### COVID-19 severity definitions based on institutional protocol

#### Mild COVID-19

Patients with baseline oxygen saturation ≥ 94% on room air without breathlessness but with other symptoms suggestive of COVID-19 such as fever, sore throat, myalgia, fatigue etc.

#### Moderate COVID-19

Patients with breathlessness with the respiratory rate (RR) ≥ 24/min and other symptoms suggestive of COVID-19 as described above, and with oxygen saturation ≤ 94% on room air.

#### Severe COVID-19

Patients with COVID-19 symptoms and RR ≥ 30 as described above with oxygen saturation ≤ 90% on room air.

### Statistical analysis

Data retrieved were cross-checked verified and entered MS Excel software version 16.0 (Microsoft Inc.). Statistical analysis was performed using SPSS Version 24 (SPSS Inc, Chicago IL, USA). The patient’s quantitative clinical data were presented as median [interquartile, IQR 25^th^ –75^th^], and categorical data were presented as numbers and proportions. Data summary was tabulated for comparison and statistical significance was checked using Mann Whitney U test and Fisher’s exact tests for quantitative and qualitative data respectively. A P-value less than 0.05 was considered significant.

## Result

### Baseline demographic and clinical characteristics of patients in both waves of COVID-19

In the first wave of COVID-19, a total of 43 patients underwent dialysis at our facility, 36 patients among them were CKD on maintenance hemodialysis, and 7 were acute kidney injury (AKI) patients. One patient was excluded from the study because of inadequate data. Among these 35 patients, 8 patients (22.9%) died during the hospital stay and 27 patients (77.1%) were discharged. In the second wave, a total of 20 patients underwent hemodialysis of which only 5 patients were CKD on maintenance dialysis while the rest 15 were AKI patients. Three of (60%) them were discharged and two (40%) didn’t survive. All the patients from both the waves who had in-hospital mortality died from the complications of COVID-19 infection.

The baseline clinico-demographic characteristics of the patients from the first wave are shown in **Table 1**. The median (IQR) age of the patients were 51 (42.5-63.5) years, ranging from 28 and 84 years with 25.7% patients were over 65 years of age. The median (IQR) age in the second wave were 62 (53-75) years, ranging from 50 to 77 years Hypertension was the most common among co-morbidities with a prevalence of 91.4%, followed by diabetes in 34.3%. The median (IQR) days between symptom onset to hospitalization was 5 (2-7.5) days. The most common presenting symptoms were shortness of breath (SOB) (62.9%), fever (54.3%), and cough (45.7%) in descending order. The severity of infection was mild in most cases (42.9%), moderate (28.6%), and severe (28.6%) in the rest. Twenty-three patients (65.7%) required oxygen supplementation at admission to the hospital. A similar clinico-demographic profile was observed in 5 patients of the second wave of COVID-19.

**Table 1:** Clinico-demographic profile with outcomes of included COVID-19 cases in the study

| Variable |  | Total<br>(N=35) | Survivors<br>(N= 27) | Mortality<br>(N= 8) | P-value |
| --- | --- | --- | --- | --- | --- |
| 1. Age (years), median [IQR] |  | 51 [42.5, 63.5] | 49 [42,55] | 70 [54,74] | 0.016 (S) |
| 2. Gender, n (%) | Male | 23 (65.7) | 16 (59.3) | 7 (87.5) | 0.139 |
|  | Female | 12 (34.2) | 11 (40.7) | 1 (12.5) |  |
| 3. Primary Kidney Disease | HTN -NP | 17 (48.6) | 13 (48.1) | 4(50.0) | 0.676 |
|  | Diabetic-NP | 10 (28.6) | 7 (25.9) | 3 (37.5) |  |
|  | PN | 8 (22.9) | 7 (25.9) | 1 (12.5) |  |
| 4. Comorbidity | Hypertensive | 32 (91.4) | 25 (92.6) | 7 (87.5) | 0.778 |
|  | Diabetic | 12 (34.3) | 8 (29.6) | 4 (50.0) |  |
|  | CAD | 5 (14.3) | 3 (11.1) | 2 (25.0) |  |
|  | Others | 11 (31.4) | 8 (29.6) | 3 (37.5) |  |
| 5. Presenting symptoms | Shortness of Breath | 22 (62.9) | 16 (59.3) | 6 (75.0) | 0.744 |
|  | Fever | 19 (54.3) | 13 (48.1) | 6 (75.0) |  |
|  | Cough | 16 (45.7) | 11 (40.7) | 5 (62.5) |  |
|  | Others | 13 (37.1) | 11 (40.7) | 2 (25.0) |  |
| 6. Duration of Symptoms before test positivity (days) |  | 5 [2,7.5] | 5 [2,10] | 3.5 [2,5] | 0.183 |
| 7. Severity of Infection at admission | Mild | 15 (42.9) | 14 (51.9) | 1 (12.5) | 0.004 (S) |
|  | Moderate | 10 (28.6) | 9 (33.3) | 1 (12.5) |  |
|  | Severe | 10 (28.6) | 4 (14.8) | 6 (75.0) |  |
| 8. Chest radiograph | Bi-lateral infiltrates | 20 (57.1) | 12 (44.4) | 8 (100) | 0.005 (S) |
| 9. SPO2: Room Air at admission |  | 97 [95,98] | 97 [95,98] | 97.5 [80,99.5] | 0.649 |
| 10. Oxygen support at Admission | Present | 23 (65.7) | 15 (55.6) | 8 (100) | 0.020 (S) |
|  | Absent | 12 (34.3) | 12 (44.4) | 0 (0) |  |
| 11. Pre-COVID dialysis duration (years) |  | 0.75 [0.5,4] | 0.5 [0.25,1] | 1 [0.5,4.2] | 0.081 |

Abbreviations: (S): statistical significance i.e., P-value less than 0.05, IQR-Interquartile range,
HTN-NP (hypertensive nephropathy), PN-primary nephropathy, CAD-coronary artery disease

### Baseline Laboratory parameters

The baseline findings in hematologic parameters were a decreased mean absolute lymphocyte counts (ALC), high neutrophil-lymphocyte ratio (NLR) and a mild decrease in platelet counts. A baseline high blood urea, and serum creatinine values were observed. The markers of inflammation, C-reactive protein (CRP), interleukin-6 (IL-6), procalcitonin (PCT), ferritin, fibrinogen and D-dimer were also high at admission. The coagulation profile showed near-normal PT, APPT, and INR. A similar trend was observed in patients from the second wave with a slightly higher baseline TLC, NLR, blood urea, and serum creatinine levels. The baseline chest X-ray (CXR) was suggestive of bilateral infiltrates in 57% and 80% of the patients in the first and second wave respectively. The median (IQR) values of baseline laboratory findings in survivors and non-survivor patients in the first wave are shown in **Table 3**.

### In-hospital course & Outcomes

After an initial admission, the severity of disease progressed in few patients subsequently requiring transfer from ward, or high dependency unit (HDU) to ICU, escalation of medical treatment, upgrade of oxygen therapy, and respiratory support in the form of non-invasive ventilation (NIV), and invasive mechanical ventilation (IMV). A total of 6 patients (17.1%) required IMV with a median (IQR) duration of 4 (0.2-4.7) days on the ventilator. In the second wave 2 out of 5 patients required IMV. All patients who required IMV during hospitalization couldn’t survive. All patients underwent hemodialysis, only one patient (2.9%) had post-dialysis hypotension and arrhythmia that was managed with appropriate pharmacological measures. In the second wave, three patients (60%) had hypotension requiring albumin and vasopressors to maintain mean blood pressure. 8 patients (22.9%) out of a total of 35 patients in the first wave and 2 out of 5 patients (40%) had in-hospital mortality. The cause for mortality observed to be the complication of the severe COVID disease leading to acute respiratory distress syndrome (ARDS) with multiorgan dysfunction syndrome (MODS) with or without refractory shock. The in-hospital management and outcomes are shown in **Table 2**.

**Table 2:**
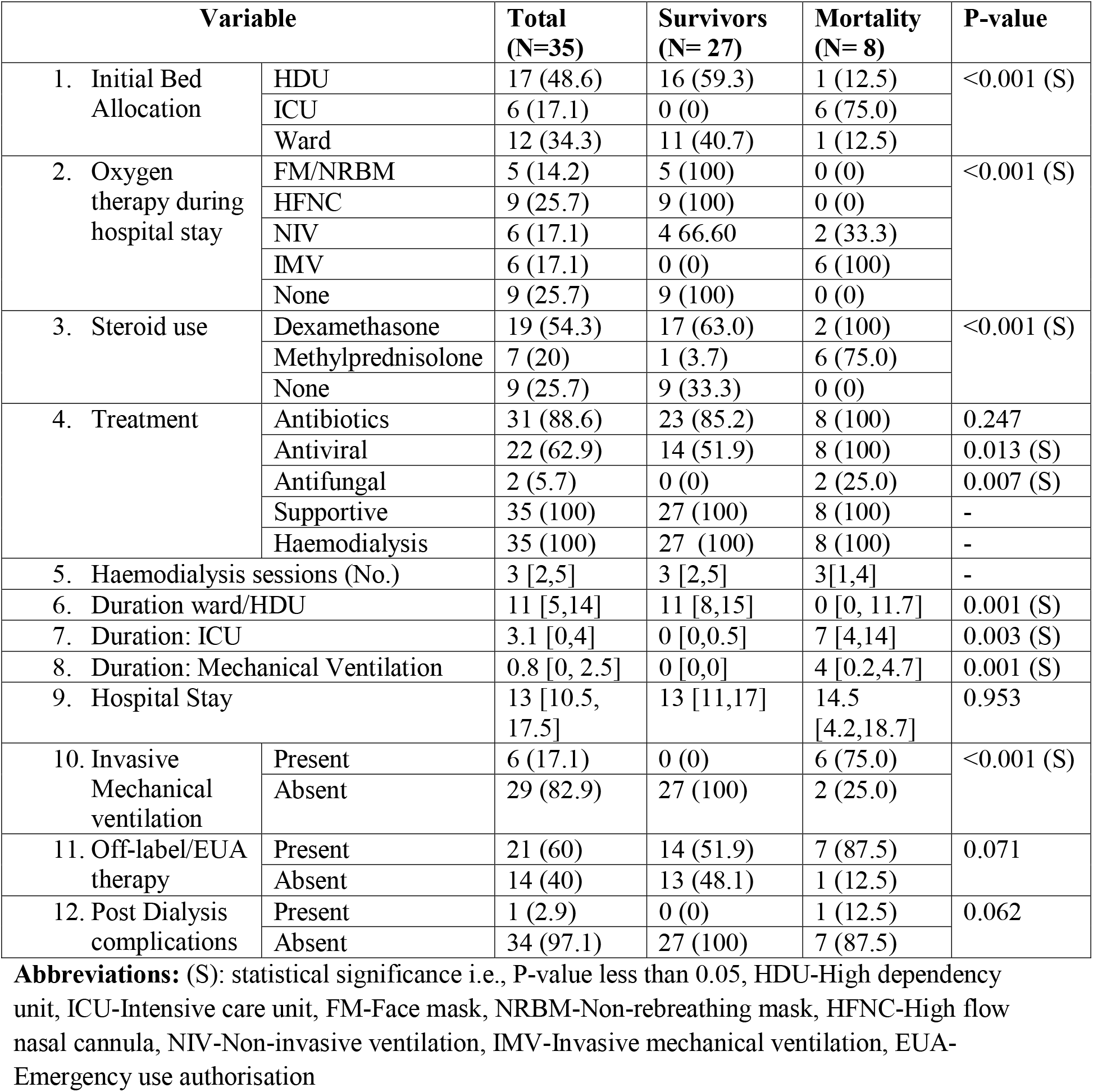
In-hospital management and outcomes of included COVID-19 cases in the study

| Variable |  | Total<br>(N=35) | Survivors<br>(N= 27) | Mortality<br>(N= 8) | P-value |
| --- | --- | --- | --- | --- | --- |
| 1. Initial Bed Allocation | HDU | 17 (48.6) | 16 (59.3) | 1 (12.5) | <0.001 (S) |
|  | ICU | 6 (17.1) | 0 (0) | 6 (75.0) |  |
|  | Ward | 12 (34.3) | 11 (40.7) | 1 (12.5) |  |
| 2. Oxygen therapy during hospital stay | FM/NRBM | 5 (14.2) | 5 (100) | 0 (0) | <0.001 (S) |
|  | HFNC | 9 (25.7) | 9 (100) | 0 (0) |  |
|  | NIV | 6 (17.1) | 4 (66.6) | 2 (33.3) |  |
|  | IMV | 6 (17.1) | 0 (0) | 6 (100) |  |
|  | None | 9 (25.7) | 9 (100) | 0 (0) |  |
| 3. Steroid use | Dexamethasone | 19 (54.3) | 17 (63.0) | 2 (100) | <0.001 (S) |
|  | Methylprednisolone | 7 (20) | 1 (3.7) | 6 (75.0) |  |
|  | None | 9 (25.7) | 9 (33.3) | 0 (0) |  |
| 4. Treatment | Antibiotics | 31 (88.6) | 23 (85.2) | 8 (100) | 0.247 |
|  | Antiviral | 22 (62.9) | 14 (51.9) | 8 (100) | 0.013 (S) |
|  | Antifungal | 2 (5.7) | 0 (0) | 2 (25.0) | 0.007 (S) |
|  | Supportive | 35 (100) | 27 (100) | 8 (100) | - |
|  | Haemodialysis | 35 (100) | 27 (100) | 8 (100) | - |
| 5. Haemodialysis sessions (No.) |  | 3 [2,5] | 3 [2,5] | 3[1,4] | - |
| 6. Duration ward/HDU |  | 11 [5,14] | 11 [8,15] | 0 [0, 11.7] | 0.001 (S) |
| 7. Duration: ICU |  | 3.1 [0,4] | 0 [0,0.5] | 7 [4,14] | 0.003 (S) |
| 8. Duration: Mechanical Ventilation |  | 0.8 [0, 2.5] | 0 [0,0] | 4 [0.2,4.7] | 0.001 (S) |
| 9. Hospital Stay |  | 13 [10.5, 17.5] | 13 [11,17] | 14.5 [4.2,18.7] | 0.953 |
| 10. Invasive Mechanical ventilation | Present | 6 (17.1) | 0 (0) | 6 (75.0) | <0.001 (S) |
|  | Absent | 29 (82.9) | 27 (100) | 2 (25.0) |  |
| 11. Off-label/EUA therapy | Present | 21 (60) | 14 (51.9) | 7 (87.5) | 0.071 |
|  | Absent | 14 (40) | 13 (48.1) | 1 (12.5) |  |
| 12. Post Dialysis complications | Present | 1 (2.9) | 0 (0) | 1 (12.5) | 0.062 |
|  | Absent | 34 (97.1) | 27 (100) | 7 (87.5) |  |

**Abbreviations:** (S): statistical significance i.e., P-value less than 0.05, HDU-High dependency unit, ICU-Intensive care unit, FM-Face mask, NRBM-Non-rebreathing mask, HFNC-High flow nasal cannula, NIV-Non-invasive ventilation, IMV-Invasive mechanical ventilation, EUA-Emergency use authorisation

### Comparison of Clinical characteristics and outcomes between survivors and non –survivors

The comparison of clinico-demographic features, in-hospital management and outcomes between survivors and non-survivors has been included in Table 1, Table 2 respectively. There was a significant difference between median (IQR) age between non-survivors versus survivors 70 (54-74) years and 49 (42-55) years respectively. The severity of COVID-19 infection at admission was also high in the non-survivor group. Seventy-five per cent (75%) of patients had the severe COVID-19 disease at admission in the non-survivor group with the presence of bilateral infiltrates on chest X-ray and requirement of oxygen and respiratory support in all patients at admission. In the survivor group, 52% of the patients had mild disease at admission with no requirement of oxygen or respiratory support in 44% of patients at admission. All the patients in the non-survivors group received higher antibiotics, antivirals, antifungals, steroids, off label and emergency use authorization (EUA) drugs following the escalation of the medical treatment. All 6 patients who required IMV during the first wave and 2 patients in the second wave didn’t survive. There were no differences in median (IQR) days of the length of hospital stay between two groups 13 (11-17) versus 14.5 (4.2-18.7) days.

### Comparison of baseline laboratory parameters between survivors and non –survivors

The comparison of baseline laboratory parameters between survivors and non-survivors has been shown in Table 3 and Figure 1. There was a significant difference in baseline Procalcitonin (PCT) and interleukin-6 (IL-6) levels with median (IQR) values of PCT 0.6 (0.2-4.4) ng/ml vs. 0.9 (0.2-9) ng/ml and IL-6 22.5 (5.7-104.6) pg/ml vs. 66.6 (10.7-184.6) pg/ml between survivors vs. non-survivors. The value of other markers of inflammation like CRP, D-dimers were also higher in non-survivors but the difference was not significant. Blood urea was significantly raised in non-survivors with a median (IQR) value of 160 (142-355) mg/dL as compared to survivors [70.6 (27.8-119.8) mg/dL]. Baseline fibrinogen and serum aspartate aminotransferase (SGOT) levels were also significantly higher, while serum albumin was significantly low in non-survivors compared to survivors [3 (2.7-3.2) g/dL versus 3.4 (3.1-3.8) g/dL] A significantly higher TLC, NLR, and lower absolute lymphocyte count has also been observed in non-survivors.

**Table 3:** Biochemical profile comparative to outcomes of included COVID-19 cases in the study

| Variable (Normal range) | Outcome |  | P-value |
| --- | --- | --- | --- |
|  | Survivors (N= 27) | Mortality (N= 8) |  |
| 1. Hb (13 - 17 g/dL) | 8.5 [7.5,10.5] | 8.9 [7.7,10.3] | 0.369 |
| 2. TLC (4 - 10 x 10<sup>3</sup>/μL) | 7.5 [5.2,12.6] | 11.4 [9.8,21.1] | 0.048 (S) |
| 3. Lymphocyte (11 – 30 x 10<sup>2</sup>/μL) | 11.1 [7,15.8] | 4.2 [3,7.6] | 0.011 (S) |
| 4. NLR (1-3) | 7.1 [5.7,12.1] | 21.8 [11.8,29.8] | 0.045 (S) |
| 5. Platelet (150 – 400 x 10<sup>3</sup>/μL) | 137 [113,276] | 135 [112,236] | 0.844 |
| 6. PT (10.2 - 13.2 sec) | 12.8 [11.6,14.2] | 12 [10.8,13.5] | 0.146 |
| 7. APTT (25.4 - 38.4 sec) | 32.8 [28.3,35.3] | 30.5 [28.5,35.3] | 0.937 |
| 8. INR (< 1.1) | 1.1 [1.0,1.2] | 1.0 [0.9,1.1] | 0.157 |
| 9. Fibrinogen (180 - 350 mg/dL) | 339 [276,449] | 392 [332,499] | 0.013 (S) |
| 10. D-dimer (< 500 ng/ml) | 330 [110.8,808.5] | 1485 [529,2856] | 0.146 |
| 11. Troponin (< 0.04 ng/ml) | 0.04 [0.01,0.09] | 0.11 [0.08,1.92] | 0.530 |
| 12. CPK (33 - 211 IU/L) | 161 [57.2,271] | 169.5 [65.5,1125.7] | 0.844 |
| 13. Ferritin (22 - 322 ng/ml) | 1204 [340,1650] | 1150 [444, 2383] | 0.252 |
| 14. CRP (0 - 0.5 mg/dL) | 9.1 [3.2,12.2] | 10.5 [3, 19.2] | 0.346 |
| 15. IL-6 (0 - 4.4 pg/mL) | 22.5 [5.7,104.6] | 66.6 [10.7,184.6] | 0.028 (S) |
| 16. Procalcitonin (< 0.1 ng/mL) | 0.6 [0.2,4.4] | 0.9 [0.2,9] | 0.037 (S) |
| 17. LDH (120 – 246 U/L) | 349.5 [254,520.2] | 453 [224,612] | 0.969 |
| 18. Urea (< 50 mg/dL) | 70.6 [27.8,119.8] | 160.1 [141.7,355.2] | 0.002 (S) |
| 19. Creatinine (0.7 - 1.3 mg/dL) | 8.5 [4.9,9.8] | 5.1 [4.1,8.5] | 0.246 |
| 20. Bilirubin (0.3 - 1.2 mg/dL) | 0.26 [0.21, 0.53] | 0.24 [0.19,0.28] | 0.270 |
| 21. SGPT (10 - 49 U/L) | 21 [9.7,35.5] | 24.3 [16.8,34.7] | 0.569 |
| 22. SGOT (< 34 U/L) | 23 [15.6,48] | 80.1 [59.7,104.7] | 0.001 (S) |
| 23. Albumin (3.2 - 4.8 g/dL) | 3.4 [3.1,3.8] | 3 [2.7,3.2] | 0.030 (S) |
| 24. Sodium (132 - 146 mmol/L) | 137 [133,139] | 135 [133,141] | 0.844 |
| 25. Potassium (3.5 - 5.5 mmol/L) | 4.1 [3.7,5.1] | 5.1 [4.3,6.1] | 0.041 (S) |
| 26. Calcium (8.7 - 10.4 mg/dL) | 7.4 [6.7,8.8] | 7.6 [7.3,7.9] | 0.906 |
| 27. Phosphorus (2.4 - 5.1 mg/dL) | 5.7 [3.9,6.7] | 3.9 [3,8.1] | 0.582 |

\* Mann Whitney U test, (S): statistical significance i.e., p value less than 0.05

**Figure 1.**
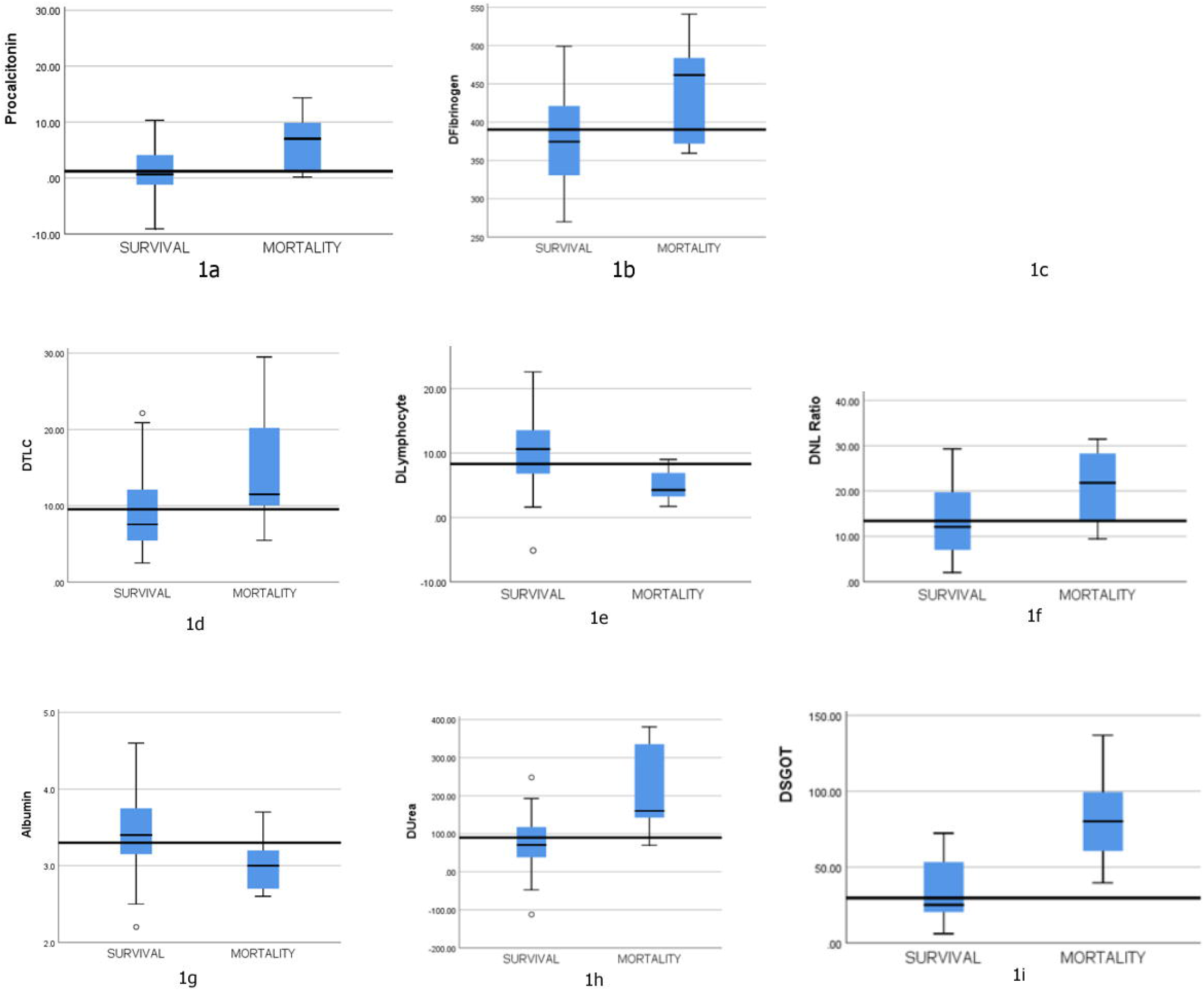
The comparison of median laboratory parameters between survivors and non-survivors, (1a) Procalcitonin, (1b) Fibrinogen, (1c) Interleukin-6 (IL-6), (1d) Total leukocyte count (TLC), (1e) Absolute Lymphocyte count, (1f) Neutrophil lymphocyte ratio (NLR), (1g) Albumin, (1h) Blood urea, (1i) Serum glutamic oxaloacetic transaminase (SGOT)

## Discussion

In this retrospective study, we aimed to determine the clinical findings, baseline laboratory parameters, and in-hospital course of CKD patients on maintenance dialysis infected with the COVID-19 virus. All patients underwent hemodialysis during the hospital stay, and concerning a modality, a majority of them received intermittent hemodialysis (IHD) technique and only a few of them underwent sustained low-efficiency daily dialysis (SLEDD). None of the patients from both the wave received continuous renal replacement therapy (CRRT). We also compared the various clinical and laboratory parameters between the survivor and the deceased group. We primarily analysed the data obtained from the 35 patients from the first wave of COVID-19 because in the second wave we had only 5 CKD patients. The number of patients undergoing dialysis for AKI (non-CKD) was high in the second wave as compared with the first wave, 68.4 % (13 out of 19 patients) versus 18.6% (8 out of 43 patients) respectively. The data obtained revealed that most patients were middle-aged, with a median (IQR) age of 51 (42.5, 63.5) years, having one or more co-morbidities along with CKD predisposing them to a high risk of severe viral infection. More than 50% of the patients (57.2%) presented with moderate to severe disease with the most reported symptom of shortness of breath (62.9%), fever (54.3%), and cough (45.7%) with a requirement of oxygen support via a variety of oxygen delivery devices at admission. Thus, the symptoms at presentation in these patients were similar to the symptoms of the general population, with no significant difference in symptomatology in survivors and non-survivors. A high baseline level of inflammatory markers like CRP, IL6, PCT, Ferritin, and LDH was observed in a majority of the patients. 75% of the patients who presented with severe disease at admission died, with 6 of them requiring invasive mechanical ventilation (IMV) during ICU stay. A significantly elevated TLC, Neutrophil Lymphocyte Ratio (NLR), IL-6, PCT, serum fibrinogen, blood urea, serum aspartate aminotransferase (SGOT), and a significantly low absolute lymphocyte count and albumin level were found in patients who had experienced unfavourable outcomes. These findings suggested that a low albumin level, coagulation abnormalities, severe inflammation, and deteriorating renal function increased the risk of mortality in these patients. A similar trend of raised inflammatory markers and high blood urea levels was observed in patients with unfavourable outcomes in the second wave, although the data were not analysed further for analysis and comparison due to the very small size of the sample (5 patients only). We observed mortality of 23% in our patients which correspond to the similar range of mortality of 16-32% reported in a similar group of patients across the various recent studies. (2–5) The complication of severe COVID-19 disease leading to acute respiratory distress syndrome (ARDS) with or without multiorgan dysfunction (MOD) was the cause of mortality in all the patients, a similar observation was reported in other previous studies as well. (6–8)

The patients with CKD on maintenance dialysis have impaired immune function. (9) The elevation in levels of inflammatory markers suggest that they exert an immunological response to the coronavirus infection and the cytokines have a key role to play in its immunopathology. (10)A high CRP level is associated with worse outcomes(2–6,11) and a similar finding in our patients supports the evidence though there was no significant elevation in non-survivors. The elevation in IL-6 and PCT were also associated with in-hospital death in infected patients. (3,12,13) Several other studies have mentioned the role of IL-6 and CRP as predictors of all-cause mortality in dialysis patients. (3,14–16) Thus, CRP, IL6 and procalcitonin levels may help to predict the progression of the severity of infection in CKD patients on maintenance dialysis. Regarding coagulation abnormalities, in our study elevated fibrinogen, D-dimer was observed in a majority of the patients, with a significant elevation of fibrinogen in non-survivors. The presence of coagulation abnormalities has been well established by previous studies in patients with COVID-19.(3,8,17) Thus, coagulation abnormalities had a higher incident rate in CKD patients on maintenance dialysis and a significant derangement is associated with poor outcomes. Other laboratory parameters like increased TLC, Neutrophil Lymphocyte Ratio (NLR), and lymphocytopenia are also associated with poor outcomes(3–6), we also observed similar findings in our study where TLC, NLR was significantly elevated in the mortality group. In routine investigations, a baseline high blood urea, SGOT, and potassium levels were associated with higher mortality in this group of patients(2,5), and similar findings were noted by us also. A low albumin level was also associated with poor outcome in these patients(2–5), that was also one of the significant findings in our study.

There are several limitations in our study, the first one being the small sample size, so the findings of the study cannot be extrapolated to the whole population of CKD patients on maintenance dialysis. Although our observations are in coherence with the findings of similar studies conducted elsewhere in the world, and a few prognostic factors are universal in this group of patients. The second limitation being the inherent information and selection bias during retrospective data retrieval. The third limitation is a very small number of CKD patients who underwent dialysis in the second wave of the COVID-19. We couldn’t compare the clinical characteristics and outcomes between the first and second wave and couldn’t come to any conclusion in this aspect. In the current literature, there are mostly retrospective studies on this group of patients so, there is a need for future similar prospective studies with an adequate sample size for better interpretation of the prognostic parameters and their association with clinical outcomes.

The observation from our study led to the conclusion that the CKD patients on maintenance hemodialysis are the group of patients that are more susceptible to severe disease following infection with coronavirus due to the presence of multiple co-morbidities, frailty, and older age. The older age, severe disease at presentation, the requirement of oxygen and respiratory support, baseline elevated inflammatory markers especially IL-6, PCT, raised fibrinogen level and a low albumin level are associated with mortality or poor outcomes in these patients.

## Data Availability

All data produced in the present study are available upon reasonable request to the authors

